# Latent Tuberculosis Diagnostics: A Systematic Review What is the past, present, and future in the diagnosis of latent tuberculosis?

**DOI:** 10.1101/2024.04.26.24306172

**Authors:** Sofia Kostoudi, Robert J H Hammond

**Author notes:** Correspondence concerning this article should be addressed to Sofia Kostoudi, School of Medical Sciences, University of Manchester, Stopford Building, Oxford Road, Manchester M13 9PL.

## Abstract

**Background:** Tuberculosis (TB) is the second leading infectious killer after COVID-19 and the 13^th^ leading cause of death worldwide. Latent tuberculosis (LTBI) has become a major pool of potential active tuberculosis cases and is propelling the TB global health burden further.

**Objective:** The creation and application of a diagnostic to effectively identify LTBI is vital. This systematic review aims to examine and analyze the present and proposed future diagnostics in the identification of latent tuberculosis.

**Design:** Systematic Review

**Methods:** PubMed and Scopus were scanned as primary databases during May 2022. Exclusion criteria for the papers scanned included patients with immunosuppression (due to HIV or treatment), pediatric TB, cancer and dialysis patients, pregnancy, IV drug users, animal models, papers published before 2005, co-infected patients, IBD and transplant patients, and finally secondary literature. Such criteria were incorporated due to the differences in TB immunology in these circumstances. 18 papers were included in this review and their risk of bias assessed using the QUADAS-2 guidelines. For analysis the papers’ sensitivities and specificities were examined. There was also a deeper look into the surrounding variables such as population differentiation, diagnostic technologies, clinical translation, and bias.

**Findings:** With thorough analysis of the data, it was determined that there are promising diagnostics for the precise identification of LTBI. Specifically, 2 studies one which used ELISA measuring the IgG response of LTBI and ATB patients when exposed to a combination of antigens and this resulted in a sensitivity and specificity of 93.33% and 93.10% respectively. The second study utilizes ESAT-6 SFC MSS (mean spot size) and the modified TBAg/PHA ratio diagnostic model to establish LTBI or ATB and using ROC curve analysis found a sensitivity of 90.12% and specificity of 91.02%.

**Interpretation:** To conclude, specific diagnostics still being examined in the preliminary phase could in the future be used as adjuncts to already present diagnostics for the diagnosis of LTBI based on their strong sensitivities and specificities.

**No funding.:** 

**SUMMARY BOX:** *What is already known on this topic?:* The WHO guidelines for diagnosing LTBI include TST and IGRA, but neither can distinguish LTBI from ATB therefore a new diagnostic must be proposed for the specific detection of LTBI

*What this study adds:* Our review reveals new two specific new diagnostic tools for the diagnosis of LTBI

*How this study might affect research, practice, or policy:* Our study can impact the future specific diagnosis of LTBI through proposing new ways of differentiating between ATB and LTBI and their possible further progression into clinical practice

## INTRODUCTION

### 1.1 Why is TB still relevant?

Tuberculosis (TB) is one of the world’s most successful human pathogens with incident cases being estimated at 10.6 million people globally in 2022, deaths at approximately 1.3 million the same year and with around ¼ of the global population harboring latent tuberculosis (LTBI) (1),(2), (3). Latent tuberculosis is defined as a state of constant immune response to *Mycobacterium tuberculosis* (M.tb) antigens. This state commonly presents without clinical manifestations of active tuberculosis disease and is noninfectious (4). The period of time over which an individual remains in a latent state of disease is variable and a healthy, immunocompetent person can harbor latent TB for their whole life 5 to 15% will go through reactivation; a process by which latent infection progresses into active TB disease (4). Therefore, individuals with LTBI can become a major reservoir for new active TB cases.

### 1.2 Importance of the Granuloma

Creating specific diagnostics for LTBI and thus identifying patients early, along with providing subsequent treatment is key when it comes to achieving the “End Tuberculosis Strategy” goal set by the WHO. At the heart of precise diagnosis for latent tuberculosis is understanding the immune response during LTBI and how this differs with active infection (ATB). Not only systemically (Table 1) with the involvement of various immune factors, but specifically in the context of the granuloma. A failure of the immune system to clear the acute infection will lead to the chronic stage characterized by the development of the granuloma. Granulomas form in approximately 90% of infected subjects and may lead to LTBI (5). The granuloma is made up of predominately macrophages CD4+, CD8+ T-cells, B cells, neutrophils, and fibroblasts (6). T-cells found in the granuloma play one of the most important roles when it comes to differentiating between LTBI and active TB disease states. In LTBI single or poly cytokine producing CD4+ T-cells are the most prevalent which secrete IFN-γ+ and IL-2+ or just IFN-γ+. (6) While in active TB different, poly functional CD4 T-cells are in abundance which secrete IFN-γ+, TNF+, and IL-2+ together (6). They are also predominantly found in the circulation of the patient rather than the granuloma (6). Activation and transitioning from LTBI to ATB via burst of the granuloma and escape of the bacteria, has been associated with various risk factors including HIV, organ transplantation, silicosis, use of TNF blockers, renal dialysis, and vitamin D deficiency (7). A low CD4+ T-cell count, an immunocompromised state, has shown to increase susceptibility to activation or reinfection which in the context of multi morbidity presents a major public health problem (6).

**Table 1:** Immune Factors.

| Immune Factor | Function Against M.tb |
| --- | --- |
| Transforming Growth Factor (TGF-β) | ☐ Blocks lymphocyte proliferation and the function of CD4 T-cells are particularly at risk (8).<br>☐ Suppresses interleukin- 2 production, suppresses expression of IFN-γ receptor and of major histocompatibility complex (MHC)-II (8).<br>☐ Blockage of IFN-γ induced macrophage activation (since it suppresses expression of IFN-γ receptors) (8).<br>☐ Capable of inducing collagen synthesis and promoting fibrosis (8). |
| Tumor Necrosis Factor Alpha (TNF-α) | ☐ Important in macrophage activation during initial contact with M.tb and cell recruitment to the site of infection (9).<br>☐ Serves as a primary signal important in granuloma formation and maintenance (9). |
| IFN-γ | ☐ Plays an important role in macrophage activation including increased pinocytosis, receptor-mediated phagocytosis, and microbial killing ability (10).<br>☐ Promotes the development of a Th1 T-cell response and it is required for a robust CD8 T-cell response (11). |
| Interleukins | ☐ Involved in the development of Th1 type of the protective immune response (12). |
| Complement Proteins | ☐ C1q is specifically associated with proinflammatory cytokine production (13).<br>☐ Can modulate dendritic cell maturation, as well as influence T- and B-cell responses (13). |

The table above provides information about key immune factors mentioned throughout the introduction that aside from many other roles they have in the body play a massive part in the immune system's defense against M.tb infection.

### 1.3 Anatomy of a Granuloma and the Role of T-cells in M.tb infection

### 1.4 Current TB Diagnostics

Effectively controlling TB requires a multifactorial approach which includes diagnostics. The three different diagnostic families used in tuberculosis detection are genotypic, phenotypic, and immunological. Genotypic tests analyze DNA (14); specific sections of the *Mycobacterium tuberculosis* genome form distinct patterns that can help differentiate strains of M.tb (14). Genotyping is primarily used to distinguish between recently transmitted and reactivation disease and to characterize evolution and phylogeny of the bacteria (15). Such information is used to gain a better understanding of pathogen-specific risk factors for transmission and pathogenesis (15).

Phenotypic tests record growth, morphology, kinetics, and metabolism.

Immunological testing involves the use of M.tb antigens to induce an antibody response. Immunological tests usually involve a skin reaction, extraction of blood, and potentially in the future, saliva (16). The biomarkers measured are from M.tb antigens directly or are a TB induced immune response and can be separated into four categories:

1. M.tb components
2. Antibody responses to M.tb antigens
3. Cellular response to M.tb antigens
4. Metabolic processes and the genetic material of M.tb (16).

The primary markers used in current TB diagnosis are antibody responses and cellular responses.

The two immunological tests used are the Tuberculin Skin Test (TST) and Interferon Gamma Release Assay (IGRA). TST uses tuberculin a purified protein derivative (PPD) to induce a type IV hypersensitivity skin reaction (17). Interferon Gamma Release Assays are *in vitro* blood tests of cell-mediated immune response and they measure T-cell release of IFN-γ after stimulation by antigens of the *M. tuberculosis* complex (18). Early secreted antigenic target 6 (ESAT-6) and culture filtrate protein 10 (CFP-10) are used as antigens (18). Which are encoded by genes located within the region of difference 1 (RD1) locus of the *M. tuberculosis* genome (18). These antigens are more specific than PPD for M.tb because they are not encoded by BCG vaccine strains or most NTM species (18). Thus, its main advantage against TST is its specificity. The main drawback of both tests is that they are unable to distinguish LTBI from ATB thus making both null diagnostics for LTBI.

### 1.5 Future Diagnostics

Drawbacks of TSTs and IGRAs primarily the limitation that both are latent diagnostics for LTBI has created a necessity for the creation of a more specific diagnostic test. Potential tests created primarily focus on immunological research. Key players are the use of more specific M.tb antigens alone or in combination with ESAT-6 or CFP-10 to distinguish LTBI from active disease. Additionally, biomarkers aside from IFN-γ that are secreted by T-cells such as IL-2 in which varying concentrations may help differentiate the two TB states such as in the state of the granuloma.

### 1.6 Aims of Review

Considering the current limitations facing TB control and elimination this review aims to evaluate current research on TST and IGRAs for the diagnosis of LTBI in immunocompetent adults and analyze future diagnostics that could potentially be of use.

## METHODS

### 2.1 Databases and Keywords

The databases used to retrieve study material were PubMed and Scopus. Through putting in the keywords “Latent tuberculosis” and “diagnostics” the amount of research papers collected was 7,708.

### 2.2 Inclusion and Exclusion Criteria

To narrow down the number of papers being used in this study and make more specific for the aims of the research, inclusion and exclusion criteria have been added (table 2).

**Table 2:**
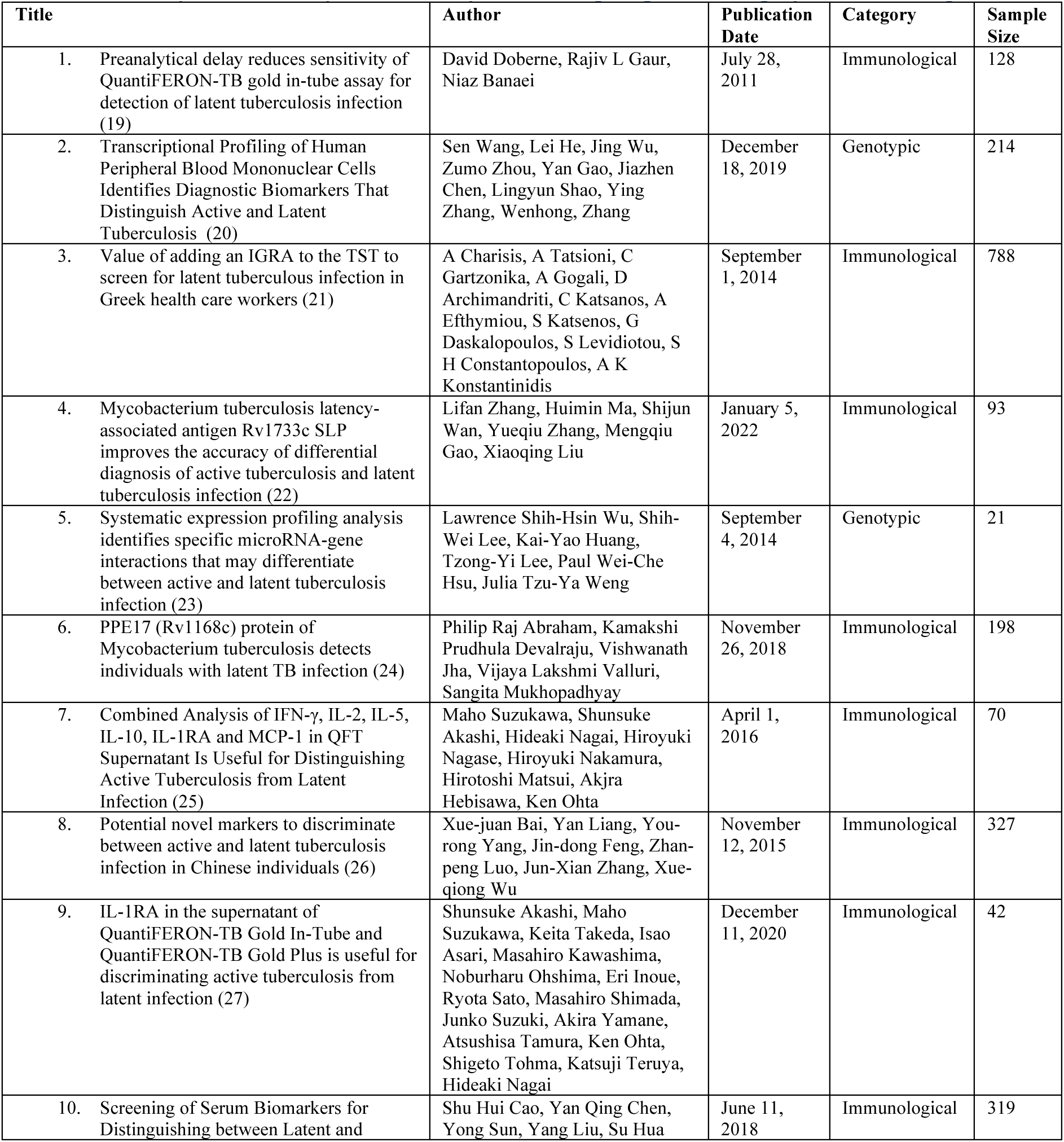
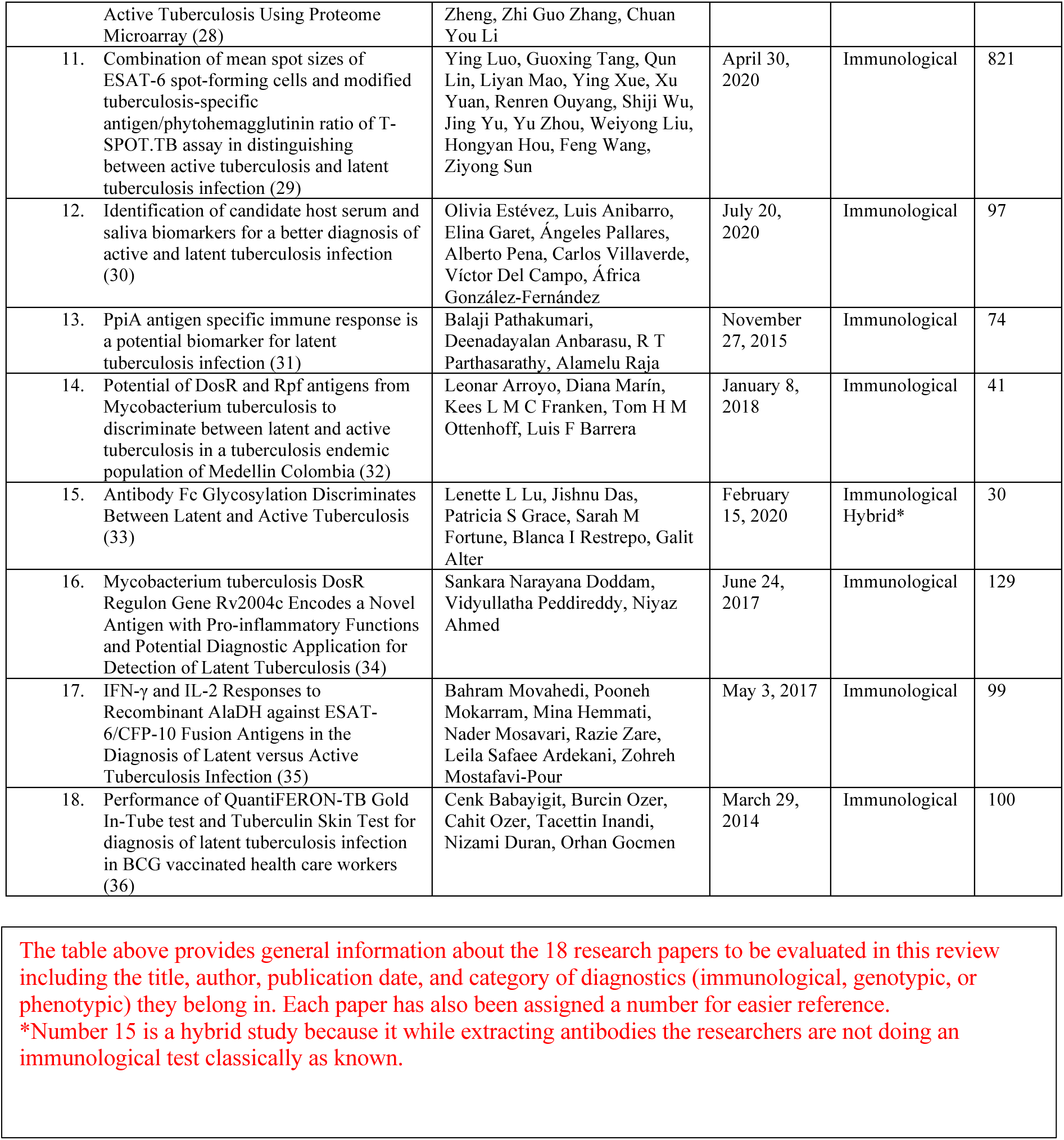
Summary of Each Study used in Analysis including diagnostic category used and sample size.

### 2.3 Research Paper Selection

The total amount of collected articles was 7,708. 1,820 duplicates were removed using EndNote software and 3 articles were retracted. 5,888 papers were screened specifically for mentions of LTBI and diagnostics as many of the collected papers were either generally about TB (pathogenesis, treatment etc.) or TB diagnostics. From this screening process a total of 905 papers were confirmed to be focused on latent tuberculosis and diagnostic mechanisms. These 905 papers were evaluated using the exclusion and inclusion criteria. A total of 386 papers remained that fit the aims and purpose of this study. Out of the 42 papers a total of 18 papers were chosen for this research (see figure 2, a PRISMA flow diagram) and their sensitivities and specificities were compared in the results along with which specific diagnostic tool (cytokine or antigen measurements etc.) each study used.

**Figure 1:**
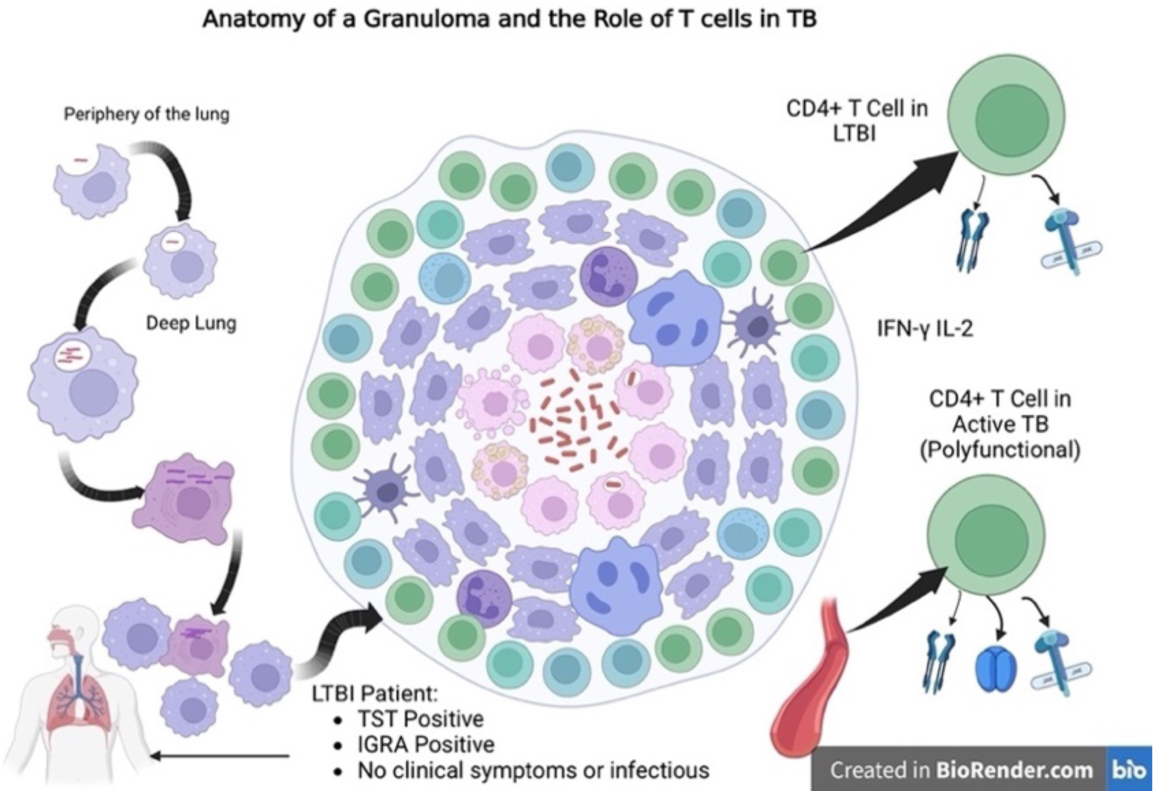
In the periphery of the lung the macrophage engulfs the bacillus specifically at the alveolar space (1). Once this occurs the macrophage moves deeper inside the lungs and the mycobacteria begin to replicate inside the macrophage. (1) The mycobacteria induce apoptosis of the macrophage via expression of MMP9 and ESX-1. These apoptotic macrophages form the core of the granuloma and newly recruited macrophages engulf the necrotic macrophages and expand the granuloma (1). Soon other cells such as T-cells arrive at the site and start to produce and release IFN-γ (1). Finally, the results the diagnostic results of an LTBI patient have been shown to complete the figure.

**Figure 2:**
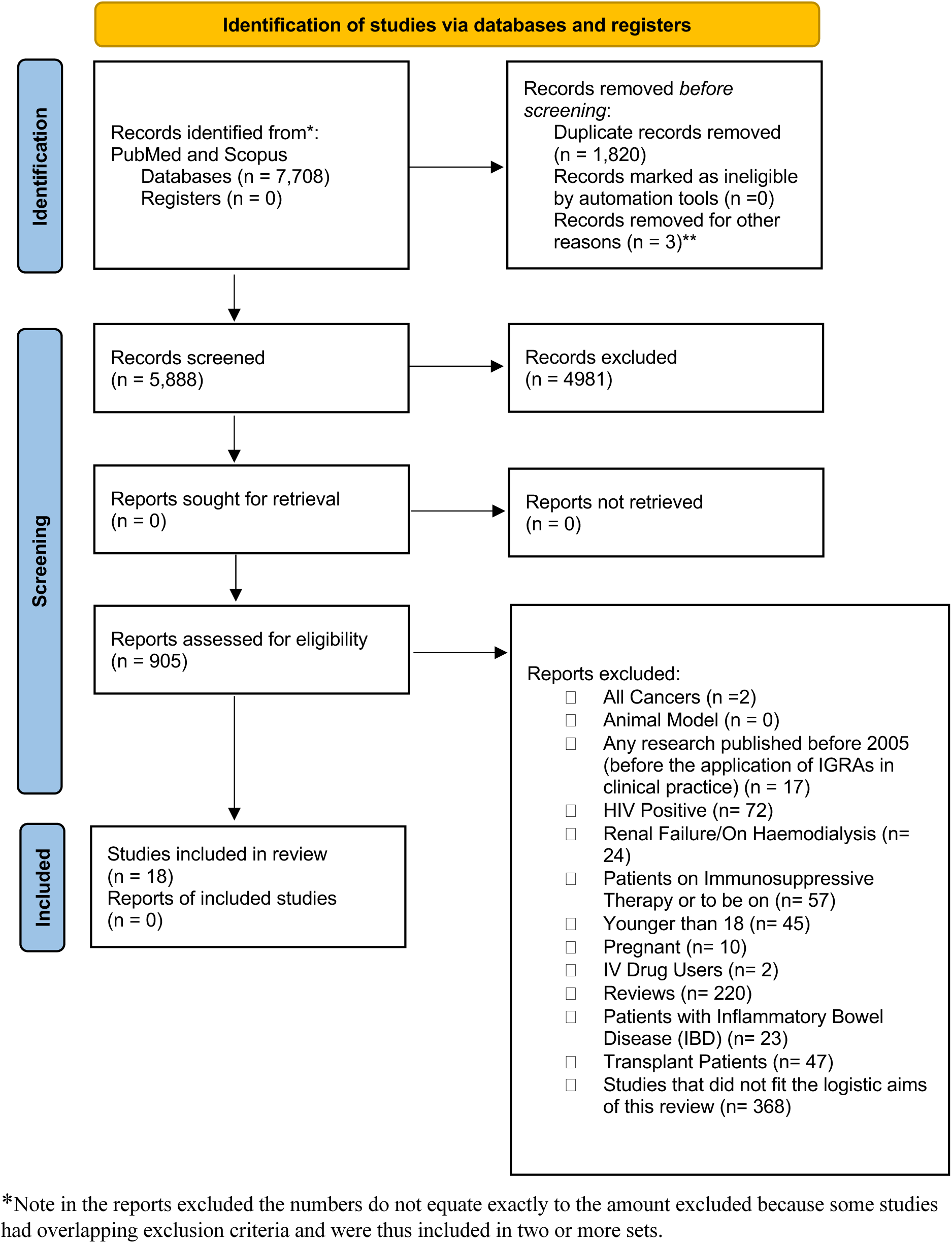
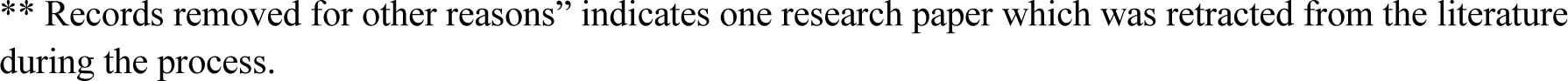
Flow diagram of the search process. PRISMA, Preferred Reporting Items for Systematic Reviews and Meta-Analyses.

**Figure 3:**
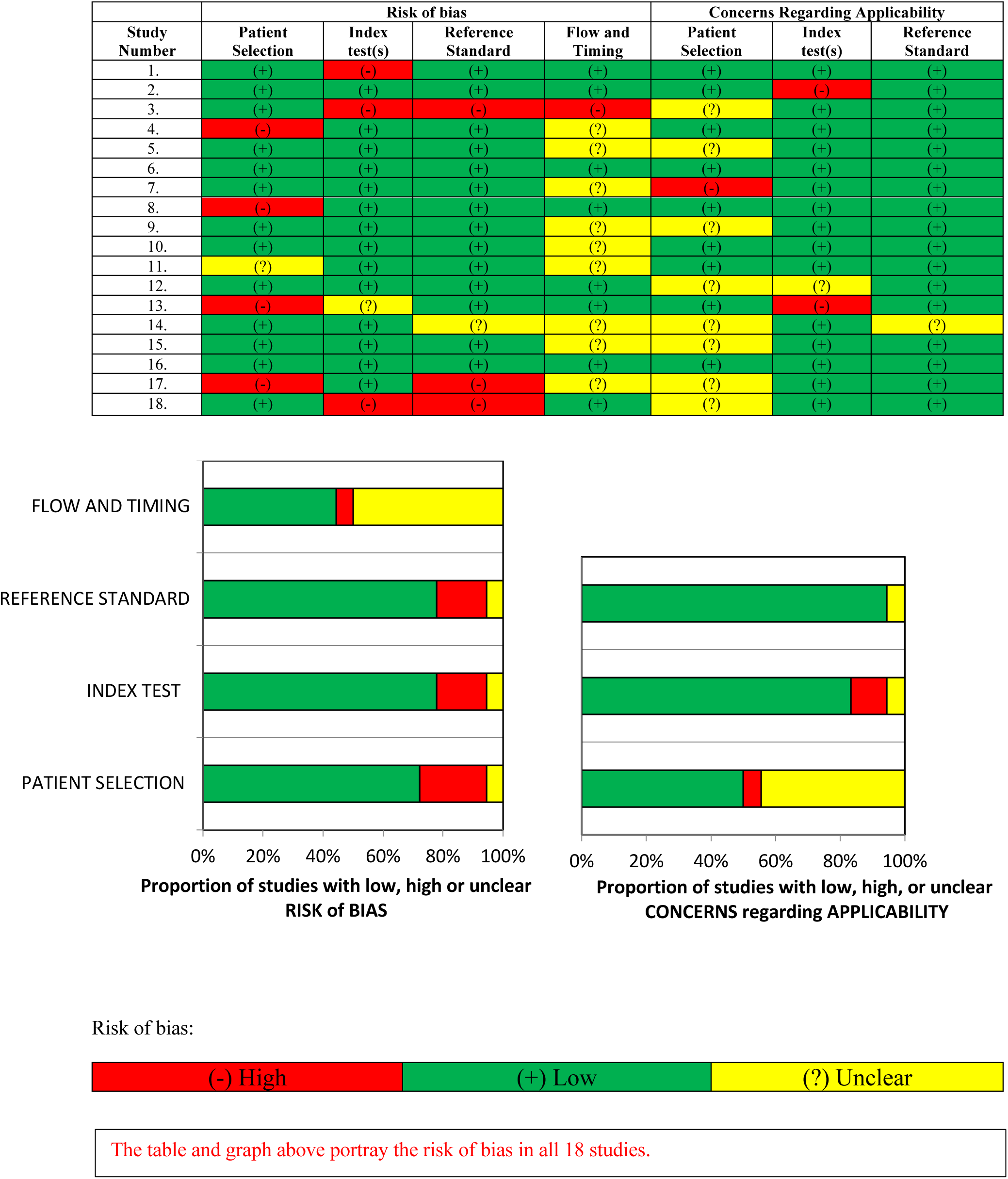
Risk of Bias Evaluated Using QUADAS-2 Guidelines:

### 2.4 Guidelines and Risk Assessment

The QUADAS-2 guidelines were used to assess the quality of the 18 studies used in terms of risk of bias, and an in-depth assessment is shown in the results section. Publication bias was reduced to the bare minimum in this review as “grey” literature was included in the research process but taken out for other reasons. The research articles were filtered using the exclusion and inclusion criteria (table 2) and not based on stronger results supporting the research question. Language bias was also removed as no articles were excluded based on language if a necessary translation was provided. Finally, no region or country was favored during the filtering process.

## RESULTS

### 3.4 Description of Results

All studies in table five have been assessed for sensitivity and specificity (if provided). Study four (22) uses FluoroSpot in testing both IFN-γ and IL-2 levels after stimulation from ESAT-6, CFP-10 separately and together or in combination with Rv1733c SLP.

Study seven (25) examined; 25 cytokines under three different conditions 11 of the 25 cytokines had promising results and could distinguish LTBI from ATB. Thus, only these 11 will be analyzed in this review.

In study eight (26) the sensitivity and specificity of different latency antigens were examined. For the first two antigens two values each are given for both sensitivity and specificity because two separate cut-off points were provided by the researchers. The reason Rv1813c does not have any values is because according to the research it had little role in differentiating between latent and active TB.

Study nine (27) examined cytokines in four conditions.

Study ten (28) analyzed 152 TB antigens and only 11 were further evaluated to determine their sensitivity and specificity in distinguishing LTBI from ATB. Finally, the antigens that performed best were studied in combination.

In study 11(29) different antigens were tested but other internal and external immune components such as MSS (mean spot size) of lymphocytes and PHA (phytohemagglutinin) were assessed. The researchers created a novel diagnostic model which combines ESAT-6 SFC MSS and modified TBAg/PHA ratio.

Study 12 (30) investigated individual host biomarkers in the serum and saliva that showed the greatest differences between groups.

Study 17 (35) : different ELISPOTs were used; one that secreted IFN-γ and another IL-2. Both measured the response of IFN-γ or IL-2 to three TB antigens ESAT-6 and CFP-10 in combination and AlaDH.

### 3.6 Risk of Bias Analysis

Risk of bias and applicability was examined for each study in general. Most studies performed well for risk of bias in patient selection, index test, and reference standard. Applicability concerns were also low for index test and reference standard. Issues for most studies presented when it came to flow and timing as well as patient selection when it came to applicability.

### 4.1 Analysis of Major Findings

Tuberculosis has become the second leading infectious killer after COVID-19 and continues to spread and propel a global health crisis (2). LTBI is a major reservoir of potential ATB where diagnostics are vital in distinguishing between the two disease states and preventing the conversion of LTBI to ATB. Each of the above 18 studies which focus on present and future LTBI diagnostics have been grouped based on the diagnostic technologies they use as this allows for more concentrated comparison.

#### Group A: Current LTBI diagnostics

Study one (19) focuses on the importance of incubation delays in generating false positive LTBI results in which the positive to negative reversion rates were 19% to 22% respectively (19). This study was conducted on healthcare workers, a key population in harboring LTBI, and highlights a strength of the work done. Such a reversion should not be present as results should be equally accurate regardless of if the sample was collected on a working day or not.

The last two studies in group A focused on the use of IGRA compared to TST in diagnosing LTBI in which both concluded that the use of IGRA is a better alternative to TST. This makes sense as IGRA utilizes much more specific antigens such as ESAT-6 and CFP-10 to mount an immune response and generate results. ESAT-6 and CFP-10 are not present in *M.bovis* of the BCG vaccine or in other mycobacterial species aside from *M.tb* allowing for a much more specific diagnosis. There are two main drawbacks in both studies though:

1. IGRA cannot distinguish between active and latent tuberculosis making it a null diagnostic.
2. Both studies were conducted in high income countries.

In the latter case resources are available to provide tests in more abundance. To fully combat TB spread, economic backgrounds of various countries must be considered. As noted by Sharma and colleagues; if current technologies cannot distinguish disease states and predict progression in the context of a resource limited and high TB burden setting TST still remains the best candidate (37). In addition, there is the advancement of TST in the context of utilizing CFP-10 and ESAT-6 but in a TST testing format, which are called Diaskintest and C-TB (38). C-TB through the clinical trials done has proven equal to IGRA and could possibly serve as a more cost effective yet equally powerful alternative to IGRA in both the high and low resource settings (39).

#### Group B: Dormancy antigens

Study four (22) examined the use of FluoroSpot with Rv1733c, ESAT-6, and CFP-10. Sensitivity and specificity were found to be 84.2% and 83.3% respectively. Study eight (26) presented similar results using just Rv2029c. Bai *et al*’s best reported T-cell responses were 84.3% in terms of sensitivity and 90% for specificity. Rv2628 yielded near identical results in terms of specificity to Rv2029c but not sensitivity which decreased to 54.3% and 48.6%. Studies 14 and 16 did not provide sensitivities and specificities immediately decreasing the apparent effectiveness and applicability of their proposed technologies. When comparing studies four and eight quantitatively study eight (26) with the usage of Rv2029c is superior. Although the above results of study eight (26) are promising Rv2029c as a potential diagnostic may have serious limitations in its use. Although it did offer high response rates in the LTBI group thus yielding the above results, it also showed greater response rates in uninfected individuals. This may be because the Rv2029c protein has homologues in other bacteria or mycobacteria. Viewed in this light Rv2029c is a weak diagnostic comparable to TST (40, 41). Rv2628 presented a moderate diagnostic potential for LTBI. In summary both dormancy antigens do not meet criteria as a strong future diagnostic specifically for LTBI.

To distinguish between LTBI and ATB the researchers used ELISASPOT and for ATB smear cultures, bronchoscopy, and chest imagining. Focusing on study four (22), the diagnostic was based on the difference of specific Th1 cytokine secretions spectrum in different TB infectious states. However, there is still an overlap in cytokine responses, which may inevitably reduce the accuracy of differential diagnosis. Study four (22) also adopted a case–control study design and probably overestimated the accuracy of the differential diagnosis as mentioned by the researchers as well. In addition, 70 Th1 cytokines have been identified in relation to tuberculosis (42). These 70 cytokines could have been investigated further by the researchers and selected in a more focused manner to which cytokines out of the 70 are more specific for LTBI (42). This would allow for a wider range of research into cytokines for the distinction of LTBI and ATB groups.

#### Group C: Transcriptional technology

RNA-seq is primarily used in identifying specific gene patterns or microRNAs to distinguish ATB from LTBI. Study two (20) utilizes the discovery of a gene that was increased in LTBI (*TNFRSF10C)* with a sensitivity of 78.4% and specificity of 84.1%. While study five (23) shows an emphasis on microRNAs which are increased in an LTBI disease state specifically hsa-miR-142-3p and hsa-miR-21-5p, but the research did not offer quantitative results for comparison highlighting a weakness of the paper. The outcomes are relatively promising in study two (20) but when examining the usage of RNA sequencing technology (RNA-seq) things are not that simple. RNA-seq has excellent potential as a diagnostic as it offers precise fine tuning of sensitivity and specificity which is key in terms of diagnostics and a wide range of reproducibility (43). Its clinical application though presents challenges namely reference standards. In order to create a diagnostic one must have reference standards representing a presence or absence of the disease state (43). With RNA seq though such a possibility is difficult as governing bodies cannot come to an agreement for reference standards or for best practice measures in the clinic on how to take RNA seq samples (43). Another potential limitation is the overabundance of software tools for sequencing technology which becomes a major obstacle in terms of standardization (43). With these two barriers it’s difficult to say when RNA-seq will become applicable for any diagnostic purposes in the clinic. Finally, because RNA sequencing techniques involve the use of genetic material, they can be very specific to a population. This can be both a pro and a con. In study two (20) the overall aims of the research differed from most of the other papers. The primary goal of paper two was to find a novel diagnostic for ATB rather than specifically LTBI. Even though a large portion of the data focused on discriminating ATB from LTBI. Such a finding presents a different approach to examine a possible diagnostic for LTBI stemming from ATB.

#### Group D: cytokines & antigens

Group D uses cytokines or antigens in identification of LTBI alone or in conjunction with IGRAs. Study six (24) provides sensitivity (65.7%) but not specificity. Study seven (25) provides a rich amount of data for diagnostic efficiency and IL-10 as the best candidate for discriminating LTBI from ATB with a sensitivity of 64.52% and specificity of 89.66% along with its high AUC. Similarly in study nine (27), IL-1RA presents the most promising quantitative results which can be seen in table 5. When comparing studies seven and nine, IL-1RA yields the most promising results from study nine (27) with the highest sensitivity and specificity, in study seven (25) coming second. In study ten (28) the combined antigen response of Rv2031c, Rv1408, and Rv2421c presented the highest sensitivity and specificity of 93.33% and 93.10% respectively. Study 12 (30) showed the combination of GRO+IL-6+IP-10+MIP1α could serve as a biomarker to distinguish LTBI from ATB with a sensitivity and specificity of 64.00% and 66.67%. Study 13 (31) has no relevant data. Study 17 (35) presented the use of antigen AlaDH in combination with CFP-10 and ESAT-6 with a response from IL-2 measured by ELISPOT where sensitivity and specificity measured 75.8% and 78.8%. A novel approach was also used to calculate the sensitivities and specificities for study 17 (35) as healthy controls were also considered calculating the above values. This is a potential strength for this study as it is equally important to be able to discriminate LTBI from healthy controls as LTBI has no clinical presentation. Out of all the studies in group D study ten (28) yielded the highest diagnostic capacity.

**Table 3:** Inclusion and Exclusion Criteria.

| Exclusion | Inclusion |
| --- | --- |
| HIV Positive | Immunocompetent adult not undergoing TB antibiotic therapy of more than 4 weeks |
| Renal Failure/On Hemodialysis | Adults (>18 yo) |
| All cancers | Lack of a history of drug use |
| Patients on Immunosuppressive Therapy or are being potentially screened for such therapy. | Data derived from primary Research |
| Younger than 18 |  |
| Pregnant |  |
| IV Drug Users |  |
| Data derived from reviews |  |
| Animal Models |  |
| Any research published before 2005 |  |
| Patients with Inflammatory Bowel Disease (IBD) |  |
| Transplant Patients |  |

**Table 4:** Sensitivity and Specificity of Current and Proposed Diagnostics for Distinguishing LTBI from ATB:

| Study Number | Sensitivity | Specificity |
| --- | --- | --- |
| 1. | NA | NA |
| 2. | 78.4% | 84.1% |
| 3. | NA | NA |
| 4. | Frequencies of single IFN-γ secreting T-cells stimulated by ESAT-6 and CFP-10: 50.9% | Frequencies of single IFN-γ secreting T-cells stimulated by ESAT-6 and CFP-10: 88.9% |
| | Proportion of single IFN-γ secreting T-cells stimulated by ESAT-6 and CFP-10: 82.5% | Proportion of single IFN-γ secreting T-cells stimulated by ESAT-6 and CFP-10: 62.2% |
|  | Frequencies of single IL-2 secreting T-cells stimulated Rv1733c SLP: 72.2% | Frequencies of single IL-2 secreting T-cells stimulated Rv1733c SLP: 73.7% |
|  | ESAT-6 and CFP-10 FluoroSpot: 82.5% | ESAT-6 and CFP-10 FluoroSpot: 66.7% |
|  | Combined Rv1733c SLP with ESAT-6 and CFP-10 FluoroSpot: 84.2% | Combined Rv1733c SLP with ESAT-6 and CFP-10 FluoroSpot: 83.3% |
| 5. | NA | NA |
| 6. | 65.57% | NA |
| 7. | IFN-γ: 90.32% | IFN-γ: 58.62 |
|  | IL-1RA: 64.52% | IL-1RA: 79.31% |
|  | IL-5: 87.1% | IL-5: 51.72% |
|  | IL-8: 66.67% | IL-8: 82.76% |
|  | IL-10: 64.52% | IL-10: 89.66% |
|  | IL-12: 54.84% | IL-12: 79.31% |
|  | IL-15: 83.87% | IL-15: 65.52% |
|  | IL-17: 87.1% | IL-17: 51.72% |
|  | MCP-1: 51.61% | MCP-1: 86.21% |
|  | PDGF-BB: 90.32% | PDGF-BB: 37.93% |
|  | RANTES: 53.33% | RANTES: 79.31% |
| 8. | Rv2029c: 84.3%, 80.4% | Rv2029c: 80%, 90% |
|  | Rv2628: 54.3%, 48.6% | Rv2628: 81%, 90% |
|  | Rv1813c: N/A | Rv1813c: N/A |
| 9. | IL-1RA: GIT-Ag: 66.67% | IL-1RA: GIT-Ag: 90.48% |
|  | TB1: 66.67% | TB1: 76.19% |
|  | TB2: 85.71% | TB2: 52.38% |
|  | Nil: 100% | Nil: 28.57% |
| | IFN-γ: GIT-Ag: 85.71% | IFN-γ: GIT-Ag: 47.62% |
|  | TB1: 52.38% | TB1: 71.43% |
|  | TB2: 28.57% | TB2: 95.24% |
|  | Nil: 42.86% | Nil: 85.71% |
|  | CXCL10/IP-10: GIT-Ag: 85.71% | CXCL10/IP-10: GIT-Ag: 52.38% |
|  | TB1: 76.19% | TB1: 57.14% |
|  | TB2: 52.38% | TB2: 85.71% |
|  | Nil: 90.48% | Nil: 47.62% |
| | CCL4/MIP-1β: GIT-Ag: 76.19% | CCL4/MIP-1β: GIT-Ag: 66.67% |
|  | TB1: 38.1% | TB1: 90.48% |
|  | TB2: 57.14% | TB2: 66.67% |
|  | Nil: 47.62% | Nil: 71.43% |
|  | CCL2/MCP-1: GIT-Ag: 61.9% | CCL2/MCP-1: GIT-Ag: 80.95% |
|  | TB1: 71.43% | TB1: 61.9% |
|  | TB2: 61.9% | TB2: 80.95% |
|  | Nil: 76.19 | Nil: 52.38% |
|  | CCL5/RANTES: GIT-Ag: 52.38% | CCL5/RANTES: GIT-Ag: 76.19% |
|  | TB1: 95.24% | TB1: 33.33% |
|  | TB2: 100% | TB2: 42.86% |
|  | Nil: 76.19% | Nil: 76.19% |
|  | PDGF-BB: GIT-Ag: 42.86% | PDGF-BB: GIT-Ag: 76.19% |
|  | TB1: 95.24% | TB1: 19.05% |
|  | TB2: 66.67% | TB2: 61.9% |
|  | Nil: 76.19% | Nil: 52.38% |
| | TNF-α: GIT-Ag: 95.24% | TNF-α: GIT-Ag: 23.81% |
|  | TB1: 90.48% | TB1: 33.33% |
|  | TB2: 80.95% | TB2: 38.1% |
|  | Nil: 90.48% | Nil: 52.38% |
| 10. | Rv2013c: 75% | Rv2013c: 84.95% |
|  | Rv1408: 75% | Rv1408: 87.10% |
|  | Rv2421c: 77.17% | Rv2421c: 82.80% |
|  | Rv2716: 66.30% | Rv2716: 97.85% |
|  | Rv2002: 73.91% | Rv2002: 83.87% |
|  | Rv2097c: 60.87% | Rv2097c: 80.65% |
|  | Rv0248c: 69.57% | Rv0248c: 73.12% |
|  | Rv2026c: 67.39% | Rv2026c: 69.89% |
|  | Rv0389: 58.70% | Rv0389: 82.80% |
|  | Rv2906c: 52.17% | Rv2906c: 89.25% |
|  | Rv2928: 42.39% | Rv2928: 92.47% |
|  | Combined Antigen Response: Rv2031c, Rv1408, and Rv2421c: 93.33% | Combined Antigen Response: Rv2031c, Rv1408, and Rv2421c: 93.10% |
| 11. | ESAT-6 SFC MSS ($\times 10^{-3}$mm<sup>2</sup>): 30.45% | ESAT-6 SFC MSS ($\times 10^{-3}$mm<sup>2</sup>): 94.92% |
|  | Modified PHA Number: 31.28% | Modified PHA Number: 91.41% |
|  | Modified ESAT-6/PHA ratio: 62.55% | Modified ESAT-6/PHA ratio: 91.80% |
|  | Modified CFP-10/PHA ratio: 67.49% | Modified CFP-10/PHA ratio: 93.36% |
|  | Modified TBaG/PHA ratio: 75.72% | Modified TBaG/PHA ratio: 91.41% |
|  | Diagnostic Model: 90.12% | Diagnostic Model: 91.02% |
| 12. | Serum: IP-10+BCA1: 72% | Serum: IP-10+BCA1: 88.00% |
|  | IP-10+BCA1+IL-7: 72% | IP-10+BCA1+IL-7: 68.42% |
| | Saliva: IP-10+IFNα2: 51.85% | Saliva: IP-10+IFNα2: 68.00% |
| | GRO+IL-6+IP-10+MIP1α: 64.00% | GRO+IL-6+IP-10+MIP1α: 66.67% |
| 13. | NA | NA |
| 14. | NA | NA |
| 15. | NA | NA |
| 16. | NA | NA |
| 17. | IFN-γ ELISPOT: ESAT-6/CFP-10: 33.3% | IFN-γ ELISPOT: ESAT-6/CFP-10: 84.8% |
|  | AlaDH: 48.5% | AlaDH: 63.6% |
|  | IL-2 ELISPOT: ESAT-6/CFP-10: 69.7% | IL-2 ELISPOT: ESAT-6/CFP-10: 42.4% |
|  | AlaDH: 75.8% | AlaDH: 78.8% |
| 18. | NA | NA |

- NA- Not applicable - Sensitivity and specificity in the context of this research are defined as the capacity of the diagnostic test to discriminate LTBI from ATB - Results in table are from each individual paper and calculated by the researchers. - The majority of the authors conducted their own analysis and used ROC analysis to do so. ROC curve analysis and AUC (area under the curve) is the preferred method of determining sensitivity and specificity in studies which choose to distinguish two disease states. Only one study (6) used patients showing antibody levels greater than or equal to OD492 (ELISA) of LTBI negative sera plus to test for sensitivity.
Where there is an N/A it means that no specificity and/or sensitivity was indicated in the study analyzed or that specificity and sensitivity were not relevant in the study's goals.

**Table 5:** Grouping of Various Studies:

| Categories | Study Numbers (Groups) |
| --- | --- |
| Current LTBI Diagnostics Used in Clinical Practice <sup>1.</sup> | A. 1., 3., 18. |
| Dormancy Survival Regulator (DosR) Regulon Encoded Antigens <sup>2.</sup> | B. 4., 8., 14., 16. |
| Transcriptional Technology <sup>3.</sup> | C. 2., 5. |
| Use of Other Antigens and Measurement of Different Cytokines <sup>4.</sup> | D. 6., 7., 9., 10., 12., 13., 17. |
| Measurement of Mean Spot Size and Antigen/Phytohemagglutinin ratio <sup>5.</sup> | E. 11. |
| Antibody Glycosylation <sup>5.</sup> | F. 15. |

The 18 studies to be analyzed in the discussion have been grouped into 6 categories based generally on the technology used for each.
1. Category involves diagnostics that are used now to diagnose LTBI such as IGRAs and TSTs. 2. Dormancy Survival Regulator Regulon Encoded Antigens are a family of tuberculosis antigens which are crucial for the prevalence of the bacteria during a latent state of infection. The studies in category B have used such antigens to induce various immune responses in patients with both ATB and LTBI. 3. Transcriptional technology studies were the 2 papers that used genotypic testing as a diagnostic method. 4. Aside from use of the basic antigens ESAT-6 and CFP-10 some of studies in category D have used other antigens to induce an immune response. Others have used the same known antigens but measured various cytokines aside from IFN-γ such as IL-2. And some in the group did both (examined new antigens and cytokines). 5. Simply studies which did not fit into any of the previous categories but also were too different from each other to be put in the same. Therefore, two separate categories were created.

When further analyzing the research papers, study six (24) presents PPE17 as a diagnostic for LTBI but it doesn’t distinguish LTBI from ATB making it a null diagnostic. In testing patient populations study nine (27) showed reversion in eight patients diagnosed with LTBI and one patient with ATB out of the 42 patients at their healthcare facilities but upon enrollment had a negative or intermediate QFT and possible shifts in disease state may reflect on the resulting cytokine levels. Study 12 (30) attempts to identify differences in saliva samples of ATB patients compared to LTBI which is a noninvasive approach to diagnosing patients and one not seen before in LTBI diagnosis.

Overall, the use of cytokines as a potential diagnostic presents with challenges in the clinical setting as there are no established cut-offs in the different cytokines measured (44). This is due to a limited understanding of normal cytokine pathways and how they react in a disease state such as tuberculosis (44). So even if cut-offs are established, they must be narrow enough to exclude or include pathology which in this case needs even further research (44). All this work must be done before the scientific community can use cytokines for effective diagnosis of LTBI.

#### Groups E and F: Unique Approaches

Groups E and F both used a novel clinical approach for the discrimination of LTBI from ATB. Study 11(29) showed a sensitivity of 90.12% and 91.12% with study 15 (33) not providing such data. Study 15 (33) uses antibody Fc glycosylation to discriminate between the two disease states. Specifically, the researchers used IgG glycosylation whereby their N-glycan structures produce great diversity which largely vary depending on the age and location of the patient population (45). Therefore, testing and results become specific to the group of patients tested and generalization becomes difficult. Tuberculosis is prevalent across the globe which doesn’t make it the most ideal diagnostic. Similarly, like for many novel technologies discussed in this paper further development is required in terms of guidelines and bioprocessing in which more rigorous and standardized methodologies must be established (45).

### 4.2 Promising Research

When holistically examining sensitivities, specificities, and biases studies ten and 11(28, 29) present the most promising results. Study ten (28) used ELISA measuring the IgG response of LTBI and ATB patients when exposed to various antigens. Furthermore, the top three antigens which performed best using AUC in distinguishing between LTBI and ATB were chosen, and a combined antigen response was performed by the researchers. This resulted in a sensitivity and specificity of 93.33% and 93.10% respectively. Study 11 (29) utilizes ESAT-6 SFC MSS (mean spot size) and the modified TBAg/PHA ratio diagnostic model to establish LTBI or ATB and using ROC curve analysis establishes a sensitivity of 90.12% and specificity of 91.02%. Thus, both studies with their increased responses could in the future be used as adjuncts to already present diagnostics for the precise diagnosis of LTBI.

### 4.3 Limitations of Review

This study has several limitations. Differences in the reference standard and biomarkers determination methods may be sources of bias. Even though QUADAS-2 was used to assess the quality and risk of bias of each study it does not fully consider all forms of bias assessed in each study. This review focuses exclusively on patients who are adults and otherwise healthy aside from being infected with LTBI, to make a diagnostic applicable clinically it should apply to most patient populations. Therefore, more research needs to be done on these diagnostic models particularly in patients co-infected with HIV, on immunosuppressive therapy and on children.

### 4.4 Conclusion

Millions of people are underdiagnosed with LTBI and it’s a vital duty of the medical community to find an accessible and accurate diagnostic for LTBI and narrow the pool of potential cases. So, we can hopefully with more progress in other areas of TB research, put an end to a disease that has been plaguing us for long enough.

## REGISTRATION

Not registered.

## Author notes

Sofia Kostoudi, School of Medical Sciences, University of Manchester, Stopford Building, Oxford Road, Manchester M13 9PL

Dr. Robert J H Hammond, Medical and Biological Sciences Building, University of St. Andrews, North Haugh, St. Andrews, Fife, KY16 9TF

## Data Availability

All data produced in the present study are available upon reasonable request to the authors.

