## Supplemental Tables for "Latent Tuberculosis Diagnostics: A Systematic Review What is the past, present, and future in the diagnosis of latent tuberculosis?"

### SUMMARY OF EACH STUDY:

| Study Design and Number | Model (Test Technology) | Description of Diagnostic | Outcome | Major Findings |
|---|---|---|---|---|
| **1.**Randomized Controlled Trial | IGRA: QuantiFERON-TB Gold in-tube assay (QFT-GIT) | Stimulate T cells with M.tb antigens and measure the amount of IFN-γ secreted by these immune cells | Measured IFN-γ levels using QFT-GIT. When 37[illegible]C incubation of QFT-GIT blood sample is delayed by 6 or 12 h there is a significant decline in IFN-γ levels produced. The decline in the TB Ag-nil value with 6- and 12-hour incubation delays resulted in positive-to-negative reversions in 19% (5/36) and 21% (5/23), respectively, of individuals with positive QFT-GIT results. | All subjects with reversions had risk factors for LTBI and therefore negative results with incubation delay were likely due to false-negative results. Therefore, immediate incubation of QFT-GIT tubes is critical for detecting T-cell responses in individuals with risk factors for LTBI and reduces intermediate results. |
| **2.** Randomized Controlled Trial | RNA-seq technique | Genome-wide transcriptional profiles of peripheral blood mononuclear cells (PBMCs) from individuals with different stages of TB infection, including active TB (ATB), LTBI, and control were determined by RNA-seq. The ratios of fold changes were determined for three pair-wise comparisons (ATB vs. LTBI, ATB vs. control, and LTBI vs. control). Differentially expressed genes in the three pair-wise comparisons were | *TNFRSF10C* was the most highly differentially expressed gene among the three groups in this study. *TNFRSF10C* levels were significantly lower in the ATB group and higher in the LTBI group than in the control group, suggesting that its expression is regulated by host cells in a stage-specific manner. | Differentially expressed genes such as *TNFRSF10C* could in the future become a valuable biomarker in differentiating LTBI from ATB and healthy controls. |

| | | | | |
|---|---|---|---|---|
| | | largely dominated by genes encoding cytokines, chemokines, and receptors under stimulation by PPD antigen of M.tb | | |
| **3.** Randomized Controlled Trial | IGRAs (QFT-GIT) and TST | IGRA: Stimulate T cells with M.tb antigens and measure the amount of IFN-γ secreted by these immune cells<br>TST: intradermal injection of 0.1 ml of PPD in the forearm and indurated area is measured 48 to 72 hours after administration | Of the enrolled healthcare workers, 36.4% were TST-positive, of whom only 14.4% were IGRA-positive. Agreement between the tests was poor (κ= 0.019; 95%CI -0.014–0.05, P= 0.355). | Due to poor overall agreement between TST and IGRA the use of IGRAs as a second step in TST-positive cases provides an appropriate tool for the diagnosis of LTBI among BCG-vaccinated healthcare workers in a low TB burden setting. |
| **4.** Case Control Study | FluoroSpot assay (ELISA) | An improvement to the traditional IGRA the FluoroSpot assay can simultaneously detect multiple cytokines at the single-cell level. | Compared the difference of cytokines secreted by ATB and LTBI groups after stimulation by MTB latency-associated antigens Rv1733c and Rv1733c SLP peptides. The obtained results revealed that Rv1733c and Rv1733c SLP could induce more T cells to produce higher levels of IL-2 in LTBI than ATB. The results of the ESAT-6 and CFP-10-Fluorospot revealed that the frequency and proportion of single IFN-γ-secreting T cells in the ATB group were significantly higher than those in the LTBI group, whereas the frequency and proportion of single IL-2-secreting T cells in the ATB group were significantly lower than those in the LTBI group. | The latency associated antigen Rv1733c SLP can be used as an alternative antigen for LTBI diagnosis based on the T cell immune reaction. The combination of ESAT-6, CFP-10, and Rv1733c SLP to induce a T cell immune reaction may prove to be a helpful diagnostic in differentiating ATB from LTBI. |
| **5.** Randomized Controlled Trial | ▪ Human OneArray v6 and Human microRN | The candidate microRNAs were analyzed for associations with the candidate genes | LTBI individuals compared to active TB patients possessed a higher level of hsa-miR-223 expression as hsa-miR-223 transcription | The levels of micro-RNA gene interactions can be used as |

| | | | | |
|---|---|---|---|---|
| | A OneArray v5<br>▪ Rosetta Resolver System software<br>▪ STRING (v9.1) | to reveal potential microRNA-gene interactions, in which decreased microRNA expression may be correlated with increased target gene expression and vice versa The genes confirmed and predicted genes were then compared with differentially expressed micro-RNA-induced upregulated and downregulated genes in TB and LTBI. | has been implicated to be induced upon the initial M.tb attack to the arrest of the infection at a latent state of disease. Also, in LTBI patients both hsa-mir-16-5p and hsa-mire-221-3p appeared to be significantly upregulated compared with TB and healthy controls. | a clinical biomarker potentially in the future for the differentiation and diagnosis of LTBI and ATB. As well as help further expand knowledge around TB disease state and progression from LTBI to ATB. |
| **6.** Randomized Controlled Trial | Enzyme-linked immunosorbent assay (ELISA test) | PPE17 (Rv1168c) protein was used to stimulate an antibody response. | LTBI individuals mounted high antibody responses against PPE17. Although it cannot distinguish LTBI from ATB. | Even if PPE17 could not distinguish between LTBI and ATB it should be considered a potential biomarker to distinguish LTBI from uninfected individuals. |
| **7.** Randomized Controlled Trial | QFT | ESAT-6, CFP-10, and TB7.7 were used as antigens to stimulate immune cells in the experimental group (TBAg). A mitogen as a positive control and without stimulation as a negative control (Nil). 25 cytokines were measured. | Several cytokines have been proposed to be able to discriminate between LTBI and active TB in the stimulated plasma. IL-10 was higher in LTBI compared to ATB. On the other hand, MCP-1 and IL-1RA are increased in active TB compared to LTBI. In the unstimulated plasma MCP-1 and IL-5 may also be good candidates in differentiating between LTBI and ATB. Lastly, IL-2 is higher in LTBI in comparison to ATB. | Overall, through this study multiple cytokines have been proposed to be able to distinguish LTBI and ATB and be of potential use in assays to diagnose LTBI. |
| **8.** Randomized Controlled Trial | Antibody ELISA assay | T cell response was measured against Rv2029c, Rv2628, and Rv1813c were determined by ELISA. | Higher immune responses to these three antigens were found in the LTBI group compared to ATB. Each antigen though responded differently. Rv1813c had no diagnostic value in differentiating LTBI from TB as their difference was very small. Rv2029c gave responses in more than 80% | These results suggest that Rv2628 protein may be one of potential candidate marker in the differentiation between LTBI persons and |

| | | | | |
|---|---|---|---|---|
| | | | LTBI, which was higher than those given by Rv2628. But Rv2029c also showed greater response rates in uninfected individuals and TB patients. Rv2628 protein showed moderate sensitivity in the LTBI group and high specificity in active TB patients and uninfected healthy subjects. | TB patients. Therefore, Rv2628 and Rv1813c (less so) can be used as biomarkers to distinguish active TB from LTBI. |
| **9.**Cross-sectional study | QFT-GIT and QFT-plus | QFT-GIT and QFT-plus tests contained either TB-specific antigens, i.e., QFT-GIT antigen (GIT Ag), TB1 antigen and TB2 antigen in QFT-plus, or a mitogen as a positive control, and without stimulation as a negative control (Nil). The levels of cytokines in the QFT supernatants were analyzed. The cytokines measured were IFN-γ, IL-1RA, IL-2, IL-5, IL-6, CXCL8/IL-8, IL-12p70, CXCL10/IP-10, CCL2/MCP-1, CCL4/MIP-1β, PDGF-BB, CCL5/RANTES, and TNF-α. | The levels of IL-1RA, IFN-γ, CXCL-10/IP-10, and CCL4/MIP-1β in QFT-GIT were clearly different between active TB and LTBI. In this study IL-1RA in QFT-GIT supernatants showed the best discrimination between LTBI from active TB. Another candidate marker in discriminating LTBI from active TB was CCL5/RANTES in the nil tube. | IL-IRA distinguished well active TB from LTBI. CCL/RANTES in the nil tube may also be a good marker in discriminating active TB from LTBI. |
| **10.** Randomized Controlled Trial | Microarray and ELISA | TST and T-SPOT (IGRA) were tested on patients with LTBI, and ATB and clinical symptoms were assessed to aid in diagnosis of each. Microarray was also performed to find candidate antigens that may help distinguish LTBI from ATB. | In this current study, the researchers screened novel serum biomarkers for discrimination between LTBI and active TB at the systemic level using an Mtb proteome microarray containing 4,262 antigens. They found that the concentrations of 152 M.tb antigen-specific IgG antibodies were higher in the active TB group than in the LTBI group. | If 152 M.tb antigen-specific IgG antibodies were higher in the active TB than in the LTBI group this could serve in the future as novel biomarkers to distinguish ATB from LTBI. |

| | | | | |
|---|---|---|---|---|
| **11.** Randomized Controlled Trial | T-SPOT. TB Assay | T-SPOT: PBMCs were isolated from ATB patients or LTBI individuals and were stimulated with ESAT-6 or CFP-10 peptides for 24 h. After stimulation, PBMCs were collected and monoclonal antibodies against the following antigens were added to the cell suspensions: CD45, CD3, CD4, CD8, and CD56. After categorizing each immune cell spot size would be calculated. Further, antigen combinations were created to potentially improve the diagnostic potential. | Comparing spot size when stimulated by ESAT-6 the ATB group had a greater number of big spots compared to the LTBI group. With CFP-10 no significant difference was established. Furthermore, combination of mean spot size of ESAT-6 with the ratio of MTB-specific antigen (TBAg) to phytohaemagglutinin (PHA) (TBAg/PHA ratio) enhanced the differential capability of distinguishing LTBI from ATB. | Mean spot size and a combination of different antigens with ESAT-6 increases the accuracy of diagnosing LTBI from ATB and vice versa. |
| **12.** Randomized Controlled Trial | Multiplex Immunoassay | Multiplex immunoassay was used to evaluate different proteins of the immune response. Several protein markers were studied in serum and saliva samples. In serum the baseline concentration of cytokines IL-6, IL-7, IP-10, TGFα, TNFα and BCA-1 were measured. And in the saliva 13 candidate biomarkers were evaluated. Combinations were also created where two cytokines were measured together. | In the serum for discriminating between LTBI and active TB patients the three markers IP-10, BCA-1 and IL-7 individually showed a good performance. These three were also combined using logistic regression. The combination of IP-10 and BCA-1 showed a better performance than these two markers alone. Adding up IL-7, however, did not improve the results. In the saliva the most promising markers to distinguish between LTBI and active TB were IFN-α2 and IP-10. Unlike, serum combination of the two saliva biomarkers did not increase accuracy for differentiating LTBI from ATB. | Potential biomarkers in the serum and saliva that could distinguish between LTBI, and ATB have been proposed in this study. Especially with saliva may serve as an effective minimally invasive process to diagnose LTBI. |
| **13.** Randomized Controlled Trial | ELISA Whole blood assay | 10 cytokines, IFN-γ, tumour necrosis factor-α (TNF-α), IL-1β, IL-2, IL-6, IL-8, IL-10, IL-12p40, IL-17 and interferon gamma- | All the tested antigens showed significantly higher IFN-γ response in LTBI patients compared to pulmonary TB (PTB) patients. PpiA-specific IFN-γ response was significantly | Overall, from the results of this study PpiA-specific IFN-γ and IFN-γ/TNF-α ratio appeared |

| | | | | |
|---|---|---|---|---|
| | | inducible protein (IP-10), and two chemokines, monocyte chemotactic protein-1 (MCP-1) and MCP-2 were measured for detection of LTBI. PpiA, ESAT-6, and CFP-10 were used as primary TB antigens | higher in LTBI patients compared to PTB. TNF-α was significantly higher in PTB compared to LTBI. IFN-γ/TNF-α ratio in response to PpiA was calculated and proved to have a high positivity (86%) for LTBI compared to PTB (18%). PpiA and CFP-10-specific IL-6 secretion was significantly higher in LTBI than PTB. But it cannot be used as a useful diagnostic because LTBI showed a positivity of 86% but in PTB the positivity was even higher therefore it cannot be used as a proposed diagnostic. IL-8, IL-12p40, and MCP-2 were concluded to not be useful as specific markers for LTBI diagnosis. | to be diagnostically useful in identifying LTBI. |
| **14.** Randomized Controlled Trial | IGRA | Three *dosR* regulon-encoded (Rv1737c, Rv2029c, Rv2628), two Resuscitation Promoting (Rv0867c and Rv2389c) antigens (Rpf) from *Mycobacterium tuberculosis*(*Mtb*), and the RD1 fusion Protein ESAT6-CFP10 (E6-C10) were used in the present study. Quantitation of IFN-γ present in the supernatants of non-stimulate and antigen stimulated PBMCs was measured. | The higher the IFNγ levels produced in response to the DosR antigen Rv2029c (PfkB), which was the antigen that best predicted disease status the higher probability of being latently infected. While individuals that produced low levels of IFNγ in response to Rv2029c displayed a higher probability of being infected with PTB. The Rpf antigens (RpfA and RpfD) induced a higher production of IFNγ in stimulated PBMCs from LTBI compared to PTB and showed a high probability to discriminate disease status. Finally, E6-C10 induced a higher production of IFNγ in stimulated PBMCs from LTBI compared to PTB and that the production of IFNγ to E6-C10 antigens showed a high probability to differentiate between LTBI and PTB, although it is not the best indicator to differentiate between the two. | Human PBMC responses to E6-C10, the M.tb DosR antigens Rv2029c and the Rpf antigens, RpfA and RpfD can discriminate latent from active TB and may be used as potential candidates for improved diagnosis of LTBI. |
| **15.** Randomized Controlled Trial | Capillary electrophoretic glycan analysis | A high-throughput capillary electrophoretic approach was employed to analyze released | The addition of glycan data with PPD antigen significantly increased classification accuracy, with the best combination resulting in a median | M.tb specific antigens and Fc glycosylation together may enhance |

| | | | | |
|---|---|---|---|---|
| | | glycans from whole antigens and isolated Fc and Fab domains from plasma-derived IgG of individuals with latent and active tuberculosis. | classification accuracy of >90% using Fc domain glycans and PPD titers in distinguishing patients with LTBI and ATB. | discriminatory potential in biomarker development for LTBI. |
| **16.** Randomized Controlled Trial | ELISA Cytokine Assays | Measures the immune response to Rv2004c a member of DosR regulon in LTBI, active TB, and healthy cohorts. | Demonstrated that antibody responses to Rv2004c were significantly higher in LTBI group followed by PTB cases, as compared to healthy controls. | Results suggest a possible use of Rv2004c as a potential serodiagnostic marker for LTBI. |
| **17.** Randomized Controlled Trial | IFN-γ and IL-2 ELISPOT Assays | IFN-γ and IL-2 ELISPOT assays were used for the quantification of the responses of the T cells to recombinant AlaDH and ESAT-6/CFP-10 antigens. | The ELISPOT assays of IFN-γ responses to AlaDH and ESAT-6/CFP-10 and IL-2 responses to ESAT-6/CFP-10 were not significantly different between LTBI and active TB, while IL-2 responses to AlaDH were different significantly. LTBI was identified with sensitivity of 75.8% and specificity of 78.8% with IL-2 responses to AlaDH. Thus IL-2 responses to AlaDH are the most sensitive and specific indicator of LTBI. | ELISPOT assay of IL-2 responses to AlaDH can detect individuals with LTBI who have recently been infected. And may serve as potential diagnostic marker in the future. |
| **18.** Randomized Controlled Trial | QFT-GIT and TST | IGRA: Stimulate T cells with M.tb antigens and measure the amount of IFN-γ secreted by these immune cells<br>TST: intradermal injection of 0.1 ml of PPD in the forearm and indurated area is measured 48 to 72 hours after administration | TST is affected by previous BCG vaccinations and number of cases with QFT-GIT positive cases is increased with the TST diameter. QFT-GIT showed discordance when compared to TST as it was negative in 17 of 32 TST positive (≥15 mm) cases. | There was slight concordance between QFT-GIT and TST when the TST result was defined as positive (diameter greater than 15 mm). Thus, QFT-GIT can be used as an alternative to TST for detection of LTBI, especially in groups with high risk of LTBI and in a population with a routine BCG |

| | | | | |
|---|---|---|---|---|
| | | | | vaccination program. |

The table above summarizes each study especially what each discovered and contributed to current and future diagnostics in TB research.

Major findings are researchers and not my own. But will be analyzed further in the discussion.

### CONFLICTS OF INTEREST:

| Study Number | Conflicts of Interest |
|---|---|
| 1. | N/A |
| 2. | The authors declare that the research was conducted in the absence of any commercial or financial relationships that could be construed as a potential conflict of interest. |
| 3. | N/A |
| 4. | None |
| 5. | The authors declare that there is no conflict of interests regarding the publication of this paper |
| 6. | N/A |
| 7. | The authors have declared that no competing interests exist. |
| 8. | The authors declare that there is no conflict of interests regarding the publication of this paper. |
| 9. | Declaration of competing interest The authors have no conflicts of interest to disclose. |
| 10. | N/A |
| 11. | Declaration of Competing Interest None declared. |
| 12. | A González-Fernández is co-promotor of the company NanoImmunoTech. The authors have no other relevant affiliations or financial involvement with any organization or entity with a financial interest in or financial conflict with the subject matter or materials discussed in the manuscript apart from those disclosed. |
| 13. | The authors declare that they have no competing interests. |
| 14. | The authors declare that they have no competing interests. |
| 15. | No reported conflicts of interest. |
| 16. | The authors declare that the research was conducted in the absence of any commercial or financial relationships that could be construed as a potential conflict of interest. |
| 17. | Conflict of Interest: None declared. |
| 18. | N/A |

The table above lists any conflicts of interests that the authors may have had that could have affected their research.
N/A= not available.
